# Multicentre validation and update of a Legionella prediction score to guide testing and treatment in community-acquired pneumonia

**DOI:** 10.64898/2026.02.25.26347092

**Authors:** Melina Bigler, Sarah Dräger, Florian Zacher, Jan Hattendorf, Daniel Mäusezahl, Werner C. Albrich, the SwissLEGIO Hospital Network

## Abstract

**Objectives:** Diagnosis of community-acquired Legionnaires’ disease (CALD) relies on microbiological testing. Routine testing in hospitalised CAP patients has low positivity rates. We externally validated a Legionella prediction score, assessed its applicability in routine care, and explored potential updates.

**Methods:** We analysed data from 196 CALD patients from 20 Swiss hospitals and 196 Legionella-negative CAP controls matched by date of diagnosis (±14 days; August 2022–March 2024). We assessed the availability of the original score predictors (fever, no/dry cough, hyponatremia, elevated CRP, elevated LDH, low platelet count) in routine care and the original score’s discriminative performance. The dataset was split into development and validation cohorts to evaluate whether simplifying modifications improved predictive performance.

**Results:** The original score showed 91% (95% CI: 86–96%) sensitivity and 35% (95% CI: 28–42%) specificity at a cut-off ≥2; LDH was infrequently measured, and platelet count was a poor predictor. The simplified *SwissLEGIO* score (fever >38°C, sodium <133 mmol/L, CRP >180 mg/L, no/dry cough, prior β-lactam therapy) maintained high sensitivity (88–92%) and showed improved specificity (46–58%) at cut-off ≥2.

**Conclusion:** The *SwissLEGIO* score is an easy-to-apply screening tool to rule out CALD in hospitalised CAP patients with scores <2 and may reduce testing by 36–52% at a CALD prevalence of 4%.

## Background

Community-acquired Legionnaires’ disease (CALD) accounts for 4-7% of community-acquired pneumonia (CAP) cases in Europe [1,2]. CALD is caused by Gram-negative, facultative intracellular *Legionella* spp., which primarily infect alveolar macrophages [3]. Effective therapy therefore requires antibiotics that reach high intracellular concentrations, such as quinolones, macrolides, or tetracyclines, whereas β-lactams are ineffective [4]. Early initiation of appropriate treatment is critical, as treatment delays are associated with increased mortality and ICU admission [4–6].

European and American guidelines adopt different strategies for managing Legionella as a possible aetiology of CAP in the hospital setting. Pan-European and Swiss/German/Austrian guidelines advise against the routine use of macrolides or quinolones for the empiric treatment of moderate CAP, favouring β-lactams monotherapy instead. Accordingly, the guidelines recommend routine Legionella testing, typically by urinary antigen test (UAT), for all hospitalised patients with at least moderate CAP [2,7,8]. By contrast, US guidelines limit Legionella testing to patients with severe CAP or those with specific epidemiological risk factors, but recommend empiric therapy with a β-lactam plus a macrolide for all hospitalised patients [9].

These differing recommendations in current guidelines reflect the ongoing debate on the optimal management of CALD in hospitalised CAP patients [10]. While routine microbiological testing for CALD allows timely and targeted treatment, and improves adherence to CAP guidelines [11], the low positivity rates of such routine testing (1.5-3%) [12–14] raise concerns regarding cost-effectiveness [9,10,15]. Conversely, selective testing strategies based on CAP severity or epidemiological risk factors have failed to reliably identify CALD patients in practice [5,12]. As an alternative approach, clinical prediction scores have been proposed [16–18]. These scores aim to identify patients at higher risk of CALD and guide decisions on microbiological testing and empiric Legionella coverage. Depending on their performance, such scores could complement laboratory diagnostics by serving as an initial step in a two-step algorithm targeting high-risk patients for microbiological testing or potentially replace such testing altogether.

The most widely used score was developed by Fiumefreddo *et al.* [18] and is referenced in the Swiss CAP guidelines [8]. The score assigns one point to each of six predictors: high fever (>39.4°C), absence of sputum (no or dry cough), hyponatremia (sodium <133 mmol/L), elevated C-reactive protein (CRP >187 mg/L), elevated lactate dehydrogenase (LDH >225 U/L), and low platelet count (<171 G/L). The score demonstrated moderate to good discrimination, with a high negative predictive value (NPV) for ruling out CALD in CAP patients with scores <2 [16,19–21]. To date, the score has not been validated in settings where its use is actively recommended in guidelines, leaving its real-world applicability uncertain. This study evaluates the score’s usability and diagnostic performance and explores potential updates using data from a representative cohort of patients with CALD from 20 Swiss hospitals and a matched control group.

## Methods

### Study population

The CALD cohort comprised patients from 15 cantonal and five university hospitals in Switzerland who were enrolled in the *SwissLEGIO* study (August 2022 and March 2024) and discharged alive after the acute CALD episode [22]. The 20 hospitals collectively reported ∼45% of all notified Legionnaires’ disease cases in Switzerland during the study period [23]. For the control group, Legionella test-negative CAP patients were matched to CALD cases by date of diagnosis (±14 days) and were recruited from the University Hospital Basel and the Cantonal Hospital St. Gallen. Both hospitals recommended universal Legionella UATs in all hospitalised CAP patients. When several matches were available, one control was randomly selected using an atomic-noise random number generator.

We reviewed electronic medical records of patients with confirmed CALD and those with Legionella test-negative CAP. Patients with CALD or Legionella test-negative CAP were eligible if they were ≥18 years old, had a clinical diagnosis of pneumonia (defined as a new infiltrate on chest imaging plus symptoms suggesting CAP such as fever, chills, cough, sputum, dyspnoea, tachypnoea, or thoracic pain) and were discharged alive after the acute episode. For CALD cases, a Legionella infection was defined according to Swiss public health criteria: positive *Legionella* spp. culture from respiratory samples or positive UAT for *L. pneumophila* or PCR detection of *Legionella* spp. in respiratory samples. Legionella test-negative CAP patients required at least one of these tests with a documented negative result. We excluded patients with suspected hospital-acquired pneumonia, travel-associated pneumonia or confirmed COVID-19, but included cases with other viral or suspected viral CAP.

The study was approved by all relevant Swiss ethics commissions (BASEC no. 2022-00880 and 2023-00097).

### Clinical and laboratory parameters

We used standardised electronic case report forms to extract data from electronic medical records. Variables included age, sex, comorbidities, timing of symptom onset and diagnosis, clinical and radiological findings at admission, indicators of disease severity (hospitalisation, length of stay, ICU admission), and information on any antibiotic prescriptions. We also collected data on inflammatory biomarkers at admission, including C-reactive protein (CRP), white blood cell counts (WBC), procalcitonin (PCT), neutrophil counts, lymphocyte counts, sodium, and lactate dehydrogenase (LDH) concentrations.

Patients were considered immunocompromised if they had any of the following conditions: treatment with steroids (≥ 7.5 mg prednisolone-equivalent/day), cytostatic/ immunosuppressive drugs (including biologicals), HIV infection with CD4 < 200/µl, neutropenia (< 0.5 g/L), history of solid organ or haematopoietic stem cell transplantation, asplenia, or primary immunodeficiency.

### Statistical analysis

#### Prediction score validation

We validated the Legionella prediction score by Fiumefreddo *et al.* [18]. For each patient, we calculated the score based on the six predictors described above. We assessed the discriminative performance of the score by plotting receiver operating characteristic (ROC) curves and calculating the area under the ROC curve (AUC). We calculated sensitivity and specificity for integer score cut-offs between 1 and 6, and estimated positive and negative predictive values (PPV, NPV) for selected cut-offs at pre-test probabilities of Legionella CAP reported in the literature (referred to as prevalence; formula in Supplementary file) [1,2]. We also re-evaluated optimal cut-offs for continuous predictors using Youden’s index.

To evaluate the score’s practical applicability, we also report the proportion of missing data for all clinical and laboratory parameters [24]. LDH was not routinely measured in clinical practice. We therefore also evaluated the predictive performance of the score when LDH was omitted.

#### Prediction score update

We evaluated whether updating the score could improve its predictive performance and simplicity of its application in routine clinical care. The dataset was randomly split by hospital site (for CALD patients who were then assigned their matched Legionella test-negative CAP control) into a development set (n = 236) for predictor selection and a validation set (n = 156) for internal validation, ensuring balanced representation of cantonal and university hospitals. Additional candidate predictors were identified through literature review and expert opinion, focusing on variables readily available at emergency department admission [5,12,18–21,25–29] (list of candidate predictors in Supplementary file). From the list of candidate predictors, we selected predictors for updating the score based on univariable and multivariable logistic regression results (assessing effect size) and plotted ROC curves (assessing balance between sensitivity and specificity) using the development dataset. To identify a new fever cut-off, we performed a classification tree analysis. We calculated the sensitivity and specificity for selected score specifications.

We internally validated the best-performing updated score using the validation set, calculating sensitivity and specificity for each score cut-off and estimating NPV and PPV for selected cut-offs and selected CALD prevalences [1,2].

#### Imputation of missing data

We assume that our data is missing at random and therefore used multiple imputation to handle missing data [24]. We applied multiple imputation by chained equations, including in the model the outcome (CALD vs Legionella-negative CAP), the six continuous predictors from the Fiumefreddo score (hyponatremia, fever, no or dry cough, LDH, CRP and platelet count), age, sex, smoking status, comorbidities (asthma, COPD, sleep apnoea, heart failure, cerebrovascular disease, kidney disease, malignancy, immunosuppression), symptom duration, clinical symptoms (dyspnoea, nausea/emesis, diarrhoea, confusion, headache, hypotension), pre-treatment with β-lactams, additional laboratory parameters (leukocytes, neutrophils, lymphocytes), and disease severity measures (non-invasive/invasive ventilation, ICU admission). We generated 40 imputed datasets and obtained pooled estimates with 95% CIs using Rubin’s rules. Within-imputation variance was estimated by bootstrapping, except for AUCs, for which we used DeLong’s method.

All analyses were performed in R (version 4.5.1, R Core Team, Vienna, Austria). For ROC curves we used the *pROC*, for multiple imputation the *mice*, for bootstrapping the *boot*, for dataset splitting the *rsample,* and for regression/decision tree analysis the *rpart* R packages.

## Results

### Patient characteristics

We included 196 CALD (Supplementary file, SFigure 1) and 196 matched Legionella test-negative CAP patients (non-LD CAP). The median age was 69 years (IQR 55-78) for CALD and 72 years (IQR 56-81) for non-LD CAP patients. Males accounted for 68.4% of CALD and 64.3% of non-LD CAP patients (Table 1).

**Table 1.** Characteristics of patients with community-acquired Legionnaire’s disease (CALD) and Legionella test-negative community-acquired pneumonia (non-LD CAP) (n (%) unless stated otherwise).

|  | CALD (n=196) | non-LD CAP (n=196) | Missing (%) |
| --- | --- | --- | --- |
| **Patient characteristics** |  |  |  |
| Male | 134 (68.4) | 126 (64.3) | 0.0 |
| Age (years) (median [IQR]) | 69 [55-78] | 72 [56-81] | 0.0 |
| Body mass index, (median [IQR]) | 25.7 [23.6-29.0] | 24.6 [21.1-28.1] | 19.4 |
| Current smoker | 83 (43.0) | 50 (32.5) | 11.5 |
| **Hospital level** |  |  |  |
| Outpatient | 7 (3.6) | 0 (0.0) | 0.0 |
| Cantonal hospital | 164 (83.7) | 130 (66.3) | 0.0 |
| University hospital | 25 (12.8) | 66 (33.7) | 0.0 |
| **Comorbidities** |  |  |  |
| Diabetes mellitus | 44 (22.6) | 47 (24.0) | 0.3 |
| Heart failure | 16 (8.2) | 38 (19.4) | 0.0 |
| Other heart diseases<sup>†</sup> | 56 (28.6) | 57 (29.1) |  |
| Asthma | 9 (4.6) | 16 (8.2) | 0.0 |
| Obstructive sleep apnoea | 15 (7.7) | 14 (7.1) | 0.0 |
| COPD | 21 (10.7) | 44 (22.4) | 0.0 |
| Cerebrovascular diseases | 16 (8.2) | 35 (17.9) | 0.0 |
| Moderate or severe chronic kidney disease | 30 (15.3) | 35 (17.9) | 0.0 |
| Malignancies | 23 (11.7) | 65 (33.2) | 0.0 |
| Immunosuppression | 38 (19.4) | 46 (23.5) | 0.0 |
| CCI (median [IQR]) | 1 [0- 3] | 3 [1-4] | 0.0 |
| **Clinical manifestation at admission** |  |  |  |
| Fever > 38°C | 135 (69.9) | 69 (35.6) | 0.0 |
| Body temperature (median [IQR]) | 38.8 [37.8-39.3] | 37.2 [36.8-38.5] | 1.3 |
| Productive cough | 41 (23.8) | 93 (53.4) | 11.7 |
| No cough or dry cough | 131 (76.2) | 81 (46.6) | 11.7 |
| Shortness of breath | 101 (51.5) | 113 (57.7) | 0.0 |
| Headache | 65 (33.2) | 41 (20.9) | 0.0 |
| Fatigue | 39 (19.9) | 27 (13.8) | 0.0 |
| Confusion | 35 (18.6) | 17 (9.7) | 7.4 |
| Diarrhoea | 36 (18.4) | 29 (14.8) | 0.0 |
| Nausea or Emesis | 23 (11.7) | 30 (15.3) | 0.0 |
| Respiratory rate ≥30/min | 36 (26.1) | 37 (32.5) | 35.7 |
| Low blood pressure <90/ ≤60 mmHg | 24 (12.4) | 50 (26.5) | 2.6 |
| **Laboratory values at admission** |  |  |  |
| CRP, mg/L (median [IQR]) | 270.5 [204.6-340.5] | 120.0 [50.3-211.3] | 0.5 |
| Procalcitonin, µg/L (median [IQR]) | 1.10 [0.36-3.64] | 0.26 [0.11-1.63] | 66.3 |
| Leukocyte count, x10<sup>9</sup>/L (median [IQR]) | 11.65 [9.15-14.92] | 12.10 [8.48-16.40] | 0.5 |
| Neutrophil count, x10<sup>9</sup>/L (median [IQR]) | 10.05 [7.41-12.67] | 9.90 [6.14-13.12] | 11.2 |
| Lymphocyte count, x10<sup>9</sup>/L (median [IQR]) | 0.76 [0.50-1.08] | 0.86 [0.60-1.41] | 11.2 |
| Platelet count, x10<sup>9</sup>/L (median [IQR]) | 206 [166-267] | 231 [174-308] | 0.5 |
| Lactate dehydrogenase, U/L (median [IQR]) | 276 [215-351] | 229 [198-287] | 34.2 |
| Sodium, mmol/L (median [IQR]) | 133 [130-135] | 136 [133-139] | 0.5 |
| Oxygen saturation <90% | 56 (29.2) | 71 (36.8) | 1.8 |
| **Disease progression and outcomes** |  |  |  |
| Symptom onset to hospitalisation (days, median [IQR]) | 4 [2- 6] | 4 [2- 8] | 8.9 |
| Pre-hospital antibiotic use | 47 (26.1) | 40 (20.6) | 4.6 |
| Monotherapy β-lactams pre-hospital use | 45 (25.0) | 25 (13.0) | 5.1 |
| Length of stay (days, median [IQR]) | 7 [5- 11] | 7 [5- 11] | 0.0 |
| ICU admission | 35 (17.9) | 40 (20.4) | 0.0 |
| Bilateral infiltrates | 35 (18.1) | 67 (35.1) | 2.0 |
| Pleural effusion | 66 (36.9) | 76 (41.8) | 7.9 |
| Non-invasive ventilation | 27 (13.8) | 27 (13.8) | 0.0 |
| Invasive ventilation | 14 (7.2) | 9 (4.6) | 0.3 |

<sup>†</sup> including arrhythmia, coronary artery disease, prior myocardial infarction, hypertensive heart disease, valvular heart disease, unspecified cardiomyopathy

Heart failure, COPD, cerebrovascular diseases, and malignancy were more common in non-LD CAP patients. Immunosuppression was reported in approximately 20% of patients in both groups. ICU admission was reported in 17.9% of CALD patients and 20.4% of non-LD CAP patients. CALD patients more frequently presented with fever and extrapulmonary symptoms, while non-LD CAP patients more often reported productive cough and hypotension. Pre-hospital β-lactam monotherapy was reported in 25.0% of CALD and 13.0% of non-LD CAP patients. Laboratory findings showed higher CRP, LDH, and procalcitonin levels in CALD patients, whereas platelet count and sodium concentrations were lower.

CALD was most frequently diagnosed by UAT (91.3% of cases). A positive PCR test was available for 20.4%, and a positive culture for 17.9% of CALD patients. In total, 23.5% of CALD patients had more than one diagnostic Legionella test that was positive. Among non-LD CAP patients, Legionella status was confirmed as negative primarily by UAT (99.5%), with negative PCR available in 1.5%. A causative pathogen was identified in 36.7% of non-LD CAP patients, most commonly *Streptococcus pneumoniae* (40.3%), *Haemophilus influenzae* (12.5%), and influenza A virus (12.5%). Co-infections were reported for 18.1% of non-LD CAP patients with an identified causative pathogen (Supplementary file, STable1). In contrast, bacterial or viral co-infections occurred in only 2.6% of CALD patients.

### Validation of the original score

All six score predictors were documented on admission in 37.2% of CALD patients and 77.6% of non-LD CAP patients. LDH was the most frequently missing parameter, absent in 57.7% of CALD patients (not routinely measured in 11 of 20 hospitals) and 10.7% of non-LD CAP patients. Documentation on sputum production was missing for approximately 10% of patients in both groups. Because LDH was frequently unavailable in medical records, we performed a secondary validation of the score excluding LDH.

The original score demonstrated moderate to good discriminative performance with an AUC of 0.78 (95% CI: 0.73-0.82). Excluding LDH from the score did not affect the AUC (0.78, 95% CI: 0.74-0.83). Among individual dichotomised predictors, elevated CRP, no or dry cough (absence of sputum), and hyponatremia had the strongest discriminative ability. By contrast, elevated LDH and reduced platelet count demonstrated poor discrimination (Supplementary file SFigure 2-3).

At the previously proposed cut-off of ≥2, the original score showed a high sensitivity (91.2%, 95%-CI: 86.2-96.2%) but limited specificity (35.3%, 95%-CI: 28.4-42.1%). When LDH was omitted, a cut-off of ≥2 yielded a sensitivity of 83.2% (95% CI: 77.7-88.6 %) and a specificity of 61.0% (95% CI: 54.0-68.0%) (Table 2).

**Table 2.** Sensitivity and specificity of the original Legionella prediction score by Fiumefreddo *et al.* (all six predictors) and when LDH was omitted (total five predictors) at different score cut-offs.

| Score | Original score |  | Score without LDH |  |
| --- | --- | --- | --- | --- |
|  | Sensitivity %<br>(95%-CI) | Specificity %<br>(95%-CI) | Sensitivity %<br>(95%-CI) | Specificity %<br>(95%-CI) |
| Score $\geq 0$ | 100.0 (100.0-100.0) | 0.0 (0.0-0.0) | 100.0 (100.0-100.0) | 0.0 (0.0-0.0) |
| Score $\geq 1$ | 99.3 (98.0-100.0) | 7.7 (3.8-11.6) | 97.3 (95.0-99.6) | 17.9 (12.4-23.4) |
| **Score $\geq 2$** | **91.2 (86.2-96.2)** | **35.3 (28.4-42.1)** | **83.2 (77.7-88.6)** | **61.0 (54.0-68.0)** |
| Score $\geq 3$ | 72.6 (65.9-79.4) | 74.0 (67.6-80.4) | 49.0 (41.8-56.2) | 89.2 (84.7-93.6) |
| Score $\geq 4$ | 39.0 (31.7-46.4) | 91.9 (88.0-95.8) | 19.3 (13.7-24.9) | 97.9 (95.9-100.0) |
| Score $\geq 5$ | 14.4 (9.1-19.7) | 99.5 (98.3-100.0) | 2.9 (0.5-5.4) | 100.0 (100.0-100.0) |
| Score $\geq 6$ | 2.1 (0.0-4.5) | 100.0 (100.0-100.0) | NA | NA |

The original score had a high NPV for ruling out CALD at scores <2. At an estimated CALD prevalence of 4%, a score cut-off <2 for ruling out CALD corresponded to an NPV of 99.0% (Table 3).

**Table 3.** Positive predictive value (PPV) and negative predictive value (NPV) for selected CALD prevalences for the original score by Fiumefreddo *et al.* (6 predictors) and when LDH was omitted (total five predictors) with a cut-off ≥2.

| Prevalence (%) | Original score, cut off $\geq 2$ | | Original score without LDH, cut off $\geq 2$ | |
| --- | --- | --- | --- | --- |
|  | PPV (95% CI) | NPV (95% CI) | PPV (95% CI) | NPV (95% CI) |
| 2 | 2.8 (2.4-3.3) | 99.5 (99.0-99.8) | 4.2 (3.3-5.3) | 99.4 (99.2-99.7) |
| 4 | 5.5 (4.8-6.5) | 99.0 (98.0-99.6) | 8.2 (6.6-10.3) | 98.9 (98.3-99.3) |
| 7 | 9.6 (8.3-11.1) | 98.2 (96.5-99.3) | 13.8 (11.3-17.2) | 98.0 (97.0-98.8) |
| 10 | 13.5 (11.8-15.6) | 97.3 (94.9-99.0) | 19.1 (15.8-23.5) | 97.0 (95.6-98.2) |

We reassessed the optimal cut-offs for the continuous predictors in the score. Overall, the original thresholds were largely consistent with our findings, supporting their validity. For fever, however, our data indicate that a lower cut-off improves discrimination between CALD and non-LD CAP patients (Supplementary file, STable 3).

### Update of the prediction score

Using the development dataset, we evaluated whether the score’s predictive performance and simplicity could be further improved. In our logistic regression analyses, elevated LDH and a reduced platelet count remained weak predictors of CALD, suggesting that they could be removed from the score. Changing the CRP threshold to >180 mg/L did not affect its predictive performance. In contrast, lowering the fever threshold to >38.0 °C (optimal cut-off according to classification tree analysis) strengthened its association with CALD compared with the original >39.4°C cut-off (Figure 1A). ROC curve analysis indicated that the adjustment of the fever cut-off primarily increased the sensitivity of the predictor, with only a modest reduction in specificity relative to the original cut-off (Supplementary file, SFigure 4).

**Figure 1.**
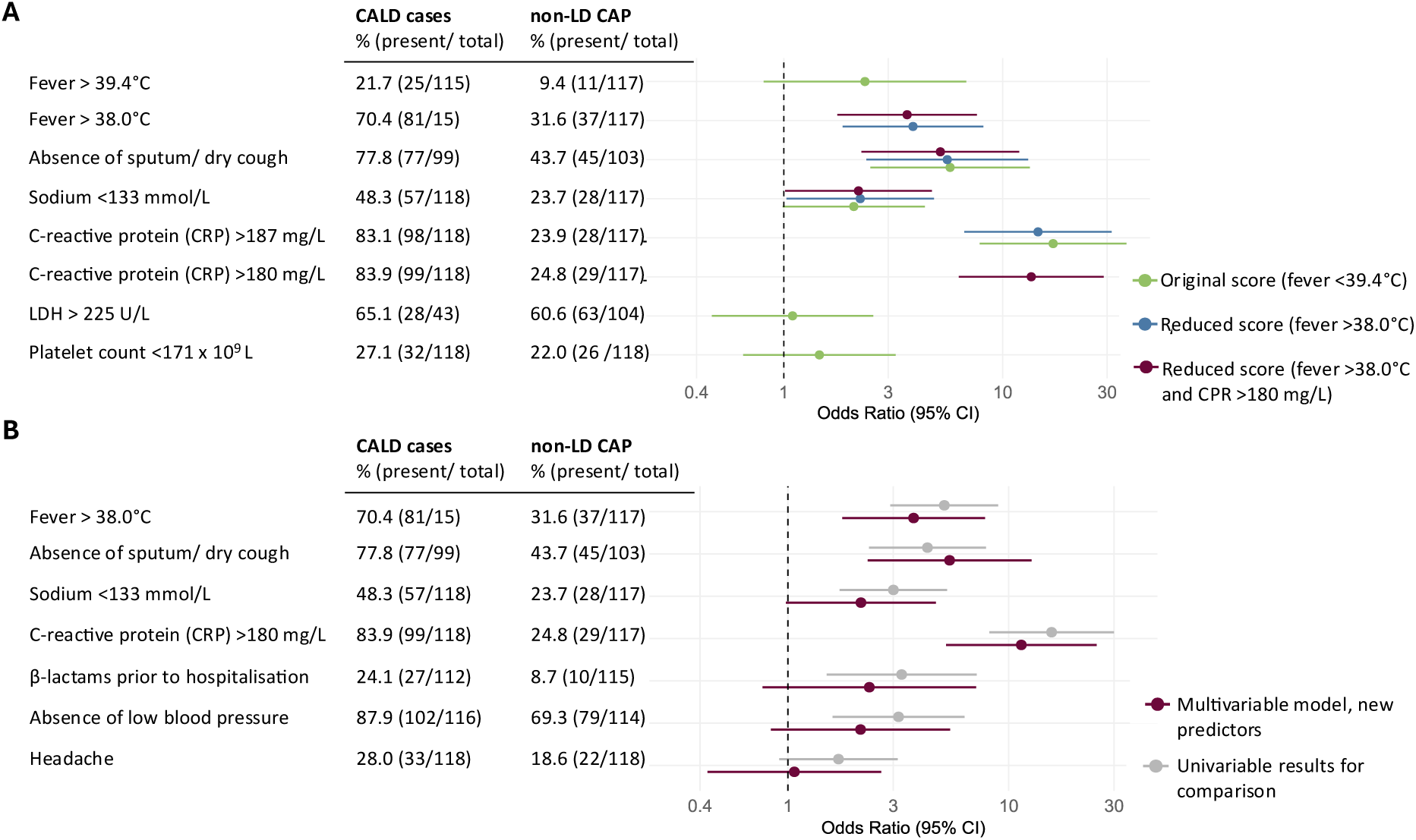
Forest plot of odds ratios (ORs) and 95% confidence intervals (CIs) from regression models fitted to imputed datasets (development dataset, n=236). Counts are based on observed data. (A) Multivariable models for the original score by Fiumefreddo *et al.*, modified 4-predictor versions with fever >38.0°C and with fever >38.0°C plus CRP >180 mg/L (B) Modified 4-predictor model with additional candidate predictors; univariable results shown for comparison.

Headache, absence of hypotension (systolic BP <90 mmHg or diastolic BP ≤60 mmHg), and prior β-lactam therapy emerged as additional predictors of CALD in the univariable logistic regression analysis (Supplementary file, SFigure 5). In the multivariable model, prior β-lactam therapy and absence of hypotension had similar effect estimates as in univariable analysis, but these associations were no longer statistically significant (Figure 1B).

We plotted ROC curves to further assess the balance between sensitivity and specificity for selected score specifications (Supplementary file, SFigure 6-7). A modified and reduced version of the score with four predictors—CRP >180 mg/L, sodium <133 mmol/L, no or dry cough (absence of sputum), and fever >38.0°C— demonstrated better predictive performance, and particularly higher specificity, than the original score by Fiumefreddo *et al*. Adding prior β-lactam therapy as an additional predictor to the reduced score (*SwissLEGIO* score, cf. Table 4) modestly increased the sensitivity at a cut-off ≥2 (Figure 2, Table 5).

**Table 4:** Simplified and revised Legionella score (*SwissLEGIO* score)

| Score predictors | Cut-off |
| --- | --- |
| No or dry cough (absence of sputum) | - |
| Fever (°C) | >38.0 |
| Sodium (Hyponatremia, mmol/L) | <133 |
| C-reactive protein (CRP, mg/L) | >180 |
| Prior β-lactam therapy | - |

One point is assigned for each parameter meeting the specified cut-off criteria. Proposed cut-off to rule-in CALD $\geq 2$ .

**Figure 2.**
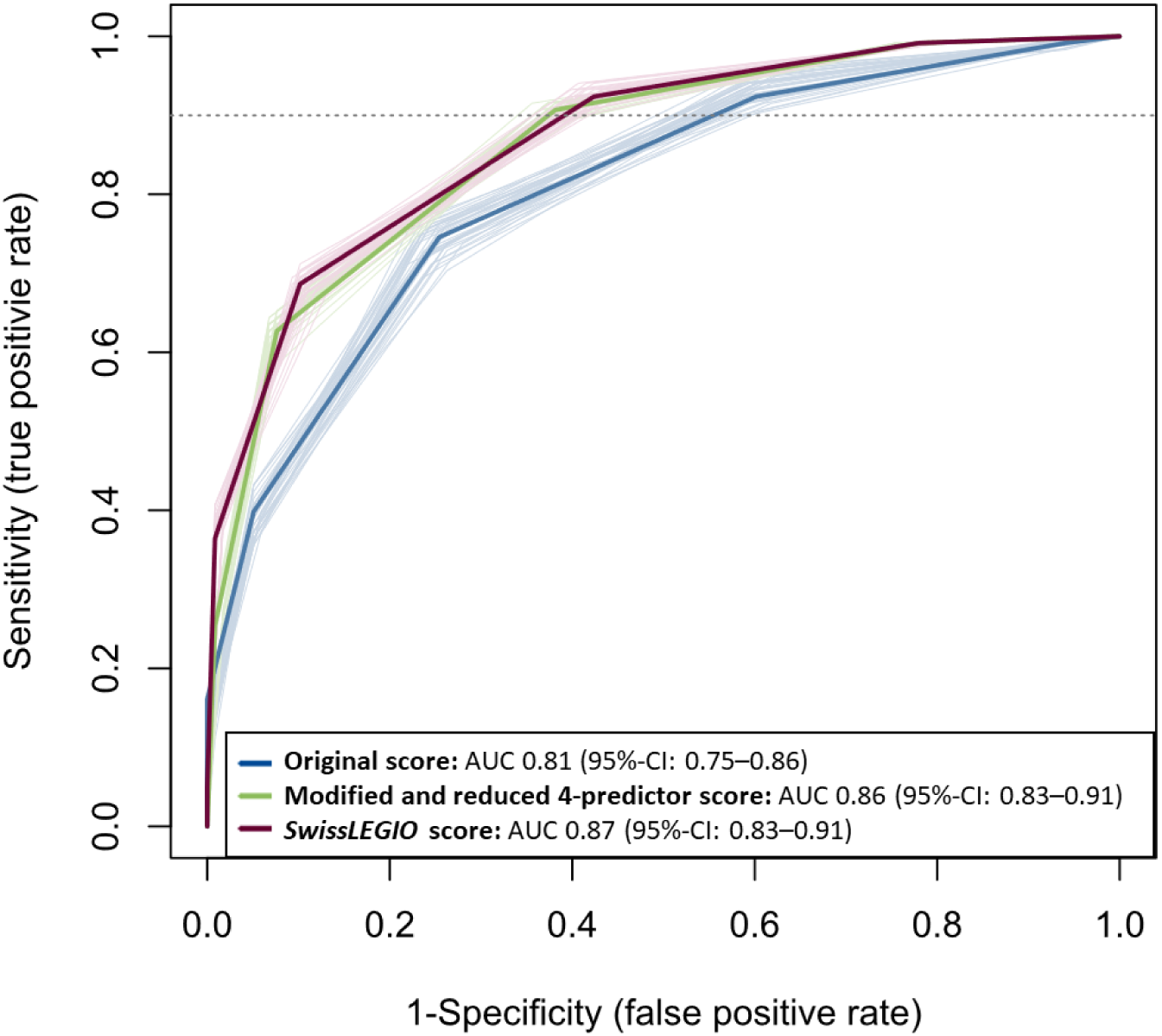
Receiver operating characteristic (ROC) curves based on the development data set (n = 236) comparing the original Fiumefreddo score, the reduced 4-predictor score (CRP > 180 mg/L, sodium, no/dry cough, fever >38 °C), and the *SwissLEGIO* score (CRP > 180 mg/L, sodium, no/dry cough, fever >38.0°C, prior β-lactam therapy). Curves from all 40 imputed datasets are shown; one is highlighted for improved readability. Dashed grey line indicates 90% sensitivity.

**Table 5:** Predictive performance of different model specifications: original Fiumefreddo score, the modified and reduced 4-predictor score (CRP > 180 mg/L, sodium, no/dry cough, fever >38 °C), and the *SwissLEGIO* score (CRP > 180 mg/, sodium, no/dry cough, fever >38.0°C, prior β-lactam therapy). CALD: community-acquired Legionnaires’ disease, AUC: area under the curve, NPV: negative predictive value; PPV: positive predictive value; CI: confidence interval. Further details are provided in Supplementary file STable 4-8.

|  | Development<br>data set (n=236) | Validation<br>data set (n=156) |
| --- | --- | --- |
| **AUC (95%CI)** |  |  |
| Original score Fiumefreddo *et al.* | 0.81 (0.75–0.86) | 0.70 (0.60–0.79) |
| Modified, reduced 4-predictor score | 0.86 (0.83–0.91) | 0.78 (0.70–0.85) |
| *SwissLEGIO* score | 0.87 (0.83–0.91) | 0.77 (0.69–0.85) |
| **Sensitivity % (95%-CI)** |  |  |
| Original score Fiumefreddo *et al.* (cut-off $\geq 2$) | 93.3 (87.7–98.8) | 86.0 (76.6–95.5) |
| Modified, reduced 4-predictor score (cut-off $\geq 2$) | 90.6 (85.1–96.1) | 85.0 (76.9–93.1) |
| *SwissLEGIO* score (cut-off $\geq 2$) | 92.3 (87.2–97.4) | 87.9 (79.9–95.9) |
| **Specificity % (95%-CI)** |  |  |
| Original score Fiumefreddo *et al.* (cut-off $\geq 2$) | 40.0 (30.7–49.3) | 28.8 (17.9–39.8) |
| Modified, reduced 4-predictor score (cut-off $\geq 2$) | 61.8 (52.7–70.9) | 49.6 (37.8–61.3) |
| *SwissLEGIO* score (cut-off $\geq 2$) | 58.1 (49.0–67.2) | 46.0 (34.0–57.9) |
| **CALD prevalence 4%: PPV % (95%-CI)** |  |  |
| Original score Fiumefreddo *et al.* (cut-off $\geq 2$) | 6.1 (5.0–7.5) | 4.8 (3.7–6.2) |
| Modified, reduced 4-predictor score (cut-off $\geq 2$) | 9.0 (7.0–12.1) | 6.6 (4.9–9.1) |
| *SwissLEGIO* score (cut-off $\geq 2$) | 8.4 (6.6–11.0) | 6.3 (4.8–8.7) |
| **CALD prevalence 4%: NPV % (95%-CI)** |  |  |
| Original score Fiumefreddo *et al.* (cut-off $\geq 2$) | 99.3 (98.4–99.9) | 98.0 (94.8–99.5) |
| Modified, reduced 4-predictor score (cut-off $\geq 2$) | 99.4 (98.8–99.8) | 98.8 (97.5–99.5) |
| *SwissLEGIO* score (cut-off $\geq 2$) | 99.5 (98.9–99.8) | 98.9 (97.6–99.7) |

In the validation data set, the modified and reduced four-predictor score and the *SwissLEGIO* score continuously showed better discriminatory ability compared to the original Fiumefreddo score. The *SwissLEGIO* score also maintained a high NPV of 98.9% for ruling out CALD in patients with a score <2 and exhibited a higher positive predictive value (PPV) of 6.3% compared to the original Fiumefreddo score (Table 5).

## Discussion

In this study, we externally validated the Legionella score developed by Fiumefreddo *et al*. and evaluated the score’s applicability in routine clinical care. The original score demonstrated moderate-to-good discriminatory performance, with high sensitivity but only moderate specificity at a cut-off ≥2. We subsequently updated the score. The simplified and revised *SwissLEGIO* score demonstrated improved discrimination, and achieved greater specificity while maintaining high sensitivity at the same cut-off (Table 4). Therefore, the updated and simplified *SwissLEGIO* score may be an effective screening tool to rule out CALD in CAP patients with a score <2.

The predictive performance of the original score in our validation cohort (sensitivity 91.2%, specificity 35.3% at a cut-off ≥2) was consistent with earlier reports. In the initial development study conducted at a Swiss university hospital, the score had an AUC of 0.86, with 92.7% sensitivity and 50.3% specificity at a cut-off ≥2. Subsequent validation studies from Europe, Japan, and the US also reported sensitivities consistently above 90%, whereas specificity ranged from 27% to 59% [16,19–21,28]. Higher specificity was mostly observed in studies with restrictive inclusion criteria, such as those requiring a confirmed CAP aetiology or complete measurement of all predictors. In contrast, studies with broader eligibility criteria reported performance estimates closer to our results.

Most of the above validation studies did not assess the feasibility of obtaining predictors in routine care. Only one Dutch study provided such data, noting that information on sputum production was missing in 31% and LDH measurements in 25% of CALD patients [20]. In our cohort, information on sputum production was missing in 10% and LDH measurements in 57.7% of CALD patients. These observations suggest that LDH is not consistently measured in routine care, likely limiting its practical usefulness as a predictor.

The predictive value of LDH for the Legionella score has also varied across studies. In the development cohort, LDH was the weakest predictor [18]. Haubitz *et al*. reported a moderate association when LDH was included as a continuous variable, but the association disappeared when LDH was dichotomised at >225 U/L [19]. Conversely, a Dutch validation study identified LDH as the strongest predictor, although this finding may have been influenced by selection bias towards patients with more severe CALD [20]. In our cohort, LDH again proved to be a weak predictor. Its predictive value may have also been partially attenuated by the presence of underlying malignancies in non-LD CAP patients. However, omitting LDH from the score improved specificity but reduced sensitivity, suggesting that LDH nonetheless contributed modestly to the score’s overall performance.

Among the remaining predictors, elevated CRP, hyponatraemia, fever, and no or dry cough (absence of sputum) were strong predictors of CALD, consistent with previous reports [12,19–21,25–30]. Platelet count was a poor predictor of CALD, a finding that was also consistently reported in earlier studies [16,19–21,28]. The predictive role of fever depended on the chosen cut-off. The original threshold of >39.4 °C offered high specificity but poor sensitivity, whereas lowering the threshold to >38.0 °C improved sensitivity with only a moderate loss of specificity. Bellew *et al*., who systematically screened hospitalised CAP patients for CALD using the UAT, previously proposed the same lower cut-off and reported an association of similar magnitude to that observed in our regression analysis [12].

Effective management of CALD is facilitated by rapid initiation of antibiotics with adequate intracellular activity, since treatment delays are associated with higher mortality and ICU admission rates [4–6]. Diagnostic scores for CALD therefore need to prioritise high sensitivity for early and reliable detection, while remaining simple to apply at hospital admission. In addition, improved specificity enhances the score’s utility in guiding microbiological testing and empiric antibiotic therapy. Accordingly, we sought to modify the original score to optimise simplicity and specificity while preserving the original score’s high sensitivity.

The updated *SwissLEGIO* score (Table 4) maintained a high sensitivity and achieved greater specificity than the original score, in both the development (40% vs. 58%) and validation data set (29% vs. 46%). However, the UAT and sputum PCR still have considerably higher specificities of 99-100% [31,32]. Therefore, the *SwissLEGIO* score should be regarded as a screening tool to guide microbiological testing, rather than as a stand-alone diagnostic method. Assuming a CALD prevalence of 4%, applying the *SwissLEGIO* score at a cut-off ≥2 would require 16 microbiological tests to identify one case, compared with 21 tests using the original score and 25 tests under universal testing. This corresponds to a 36% reduction in testing when the *SwissLEGIO* score is used for screening. In the development dataset, the reduction in testing even reached 52% when the *SwissLEGIO* score was applied instead of universal testing.

The *SwissLEGIO* score may be particularly useful in settings where routine CALD testing is not recommended. For instance, US guidelines restrict testing to patients with severe CAP or specific epidemiologic risk factors [9], but such selective testing is often inefficient [5,12], and epidemiologic risk factors can vary by setting [22]. To compensate, US guidelines adopt a lower threshold for universal combination therapy, which European guidelines generally avoid [2,7,8]. Distinguishing CALD from other CAP aetiologies based on individual symptoms is also challenging, since extrapulmonary manifestations suggestive of CALD (such as headache, confusion, gastrointestinal symptoms, or acute kidney injury) are present in fewer than half of CALD patients [26,27,29,33].

In contrast to considering individual risk factors and clinical characteristics, combining them in the *SwissLEGIO* score reliably identified CAP patients at higher risk of CALD. The score provides clear and pragmatic criteria to guide empiric antibiotic treatment and diagnostic testing decisions. Nonetheless, guideline authors should carefully weigh the efforts of implementing the *SwissLEGIO* score and the potential risk of missed cases against the score’s potential to reduce costs for microbiological testing. In settings with higher CALD incidences, routine testing with the UAT or molecular assays may therefore still be the most straightforward approach to appropriately managing CALD in the hospital setting.

Our study has limitations. First, because CALD is relatively rare, we used a case-control design with cases and controls not always drawn from the same hospital. This design is more prone to selection bias than cohort studies and does not allow direct estimation of CALD prevalences [24]. However, both hospitals enrolling controls routinely performed Legionella UATs in all CAP patients, and the characteristics of our control cohort were consistent with other studies (Supplementary file) [12,14,19]. We also excluded patients with travel-associated CALD, which were part of the initial publication by Fiumefreddo and colleagues. Second, screening for CALD relied mainly on UAT, which has the highest sensitivity for *L. pneumophila* serogroup 1 [4]. The predictive performance of the *SwissLEGIO* score might therefore vary for infections caused by other serogroups or *Legionella* species [34]. Nonetheless, in a sensitivity analysis limited to UAT-negative CALD cases, the *SwissLEGIO* score still achieved a sensitivity of 83.8%. Finally, although findings were consistent across development and validation datasets, the score has not been externally validated; we strongly encourage further external validation before broader adoption in guidelines.

## Conclusion

The updated and simplified *SwissLEGIO* score (CRP >180 mg/L, sodium <133 mmol/L, no cough or dry cough, fever >38.0°C, and prior β-lactam therapy), is an easy-to-apply screening tool to rule out CALD in hospitalised CAP patients with a score <2. The *SwissLEGIO* score can complement microbiological testing by providing clear and pragmatic criteria to decide who should undergo diagnostic testing for Legionella. The score may be particularly useful in settings where routine testing is not recommended, providing a more efficient alternative to epidemiological risk factor- or severity-guided testing.

## Supporting information

preprint_supplement_swisslegio_score

## Additional information

## CRediT authorship contribution statement

Conceptualisation of the study: M.B., S.D., W.C.A. and D.M. Data collection/ Investigation: F.Z., M.B. and the members of the SwissLEGIO hospital network. Statistical analysis: M.B with J.H. Data interpretation: M.B., S.D., W.C.A., F.Z., and D.M. Writing of the original draft of the manuscript: M.B. All authors critically revised the manuscript for important intellectual context and edited the manuscript. All the authors reviewed and approved the final version of the manuscript.

## Acknowledgment

At Swiss TPH, warmly thank Fabienne B. Fischer and Manuel Wiederkehr for their scientific input and support in coordinating the study. We further thank Gabriel Escher and Dominik Schnyder for their support during the data collection. At the Federal Office of Public Health, we thank Marianne Jost, Mirjam Mäusezahl-Feuz, and Sabine Basler for inputs and discussions surrounding the surveillance of Legionnaires’ disease. Finally, we would like to thank all the additional staff from the SwissLEGIO hospital network for their unfailing support during the data collection: Sheila Barbarossa (NRCL); Dr. Valentin Gisler and Eva Hitz (Cantonal Hospital Aarau), Camille Schilt and Dr. Andréa Künzli (Cantonal Hospital Neuchâtel); Jane Frangi and Luisa Vicari (Ente Ospedaliero Cantonale); Vanessa Deggim-Messmer and Dr. Véronique Erard (Cantonal Hospital Fribourg); Dr. Alexis Dumoulin (Valais Hospitals); Dr. Brigitte Suter Buser (Cantonal Hospital Luzern); Daniela Hirter and Pia Scherler (Insel University Hospital Bern); Dr. Peter M. Keller and Prof. Michael Osthoff (University Hospital Basel); Kristin Abig and Andrea Kloetzer, and Dr. Carmen Volken (Cantonal Hospital Baselland); Dr. Emily West (University Hospital Zurich); as well as the many additional contributors across the SwissLEGIO hospital network who supported case identification and logistics.

Declaration of the use of generative AI and AI-assisted technologies: During the preparation of this manuscript, the author(s) used Grammarly and ChatGPT (OpenAI) to assist with grammar correction and improving readability. All changes suggested by these tools were carefully reviewed and edited, and the author(s) take full responsibility for the content of the published article.

## Data availability statement

Study data will be available for research purposes upon reasonable request, respecting all relevant regulations and with the establishment of confidential data transfer agreements.

