## Supplementary material for "Multicentre validation and update of a Legionella prediction score to guide testing and treatment in community-acquired pneumonia": preprint_supplement_swisslegio_score

**Affiliations**
[a] Swiss Tropical and Public Health Institute, Kreuzstrasse 2, 4123 Allschwil, Switzerland
[b] University of Basel, Petersgraben 35, 4001 Basel, Switzerland
[c] Division of Internal Medicine, University Hospital Basel, Petersgraben 4, 4031 Basel, Switzerland
[d] Department of Clinical Research, University of Basel, Petersgraben 35, 4001 Basel, Switzerland
[e] Division of Infectious Diseases, Infection Prevention and Travel Medicine, Cantonal Hospital St. Gallen, HOCH Health Ostschweiz, Rorschacher Str. 95, 9007 St.Gallen, Switzerland

Preprint

#### Study summary

In this study, we externally validated the Legionella score proposed by Fiumefreddo *et al.* in a representative cohort of CALD patients from 15 cantonal and five university hospitals in Switzerland, along with an appropriate control group of Legionella test-negative community-acquired pneumonia (CAP) patients enrolled from one university and one cantonal hospital. CALD patients were enrolled through the *SwissLEGIO* study. We additionally assessed whether the predictive performance and simplicity of the score could be improved. To do so, we split the dataset into a development cohort (n=236) and a validation cohort (n=156). Based on our analysis, we propose a new, simplified version of the score (***SwissLEGIO* score**), comprising five easily measurable variables: **fever >38°C, sodium <133 mmol/L, CRP >180 mg/L, absent or dry cough (no sputum), and prior β-lactam therapy**.

### Additional study results: Patient characteristics

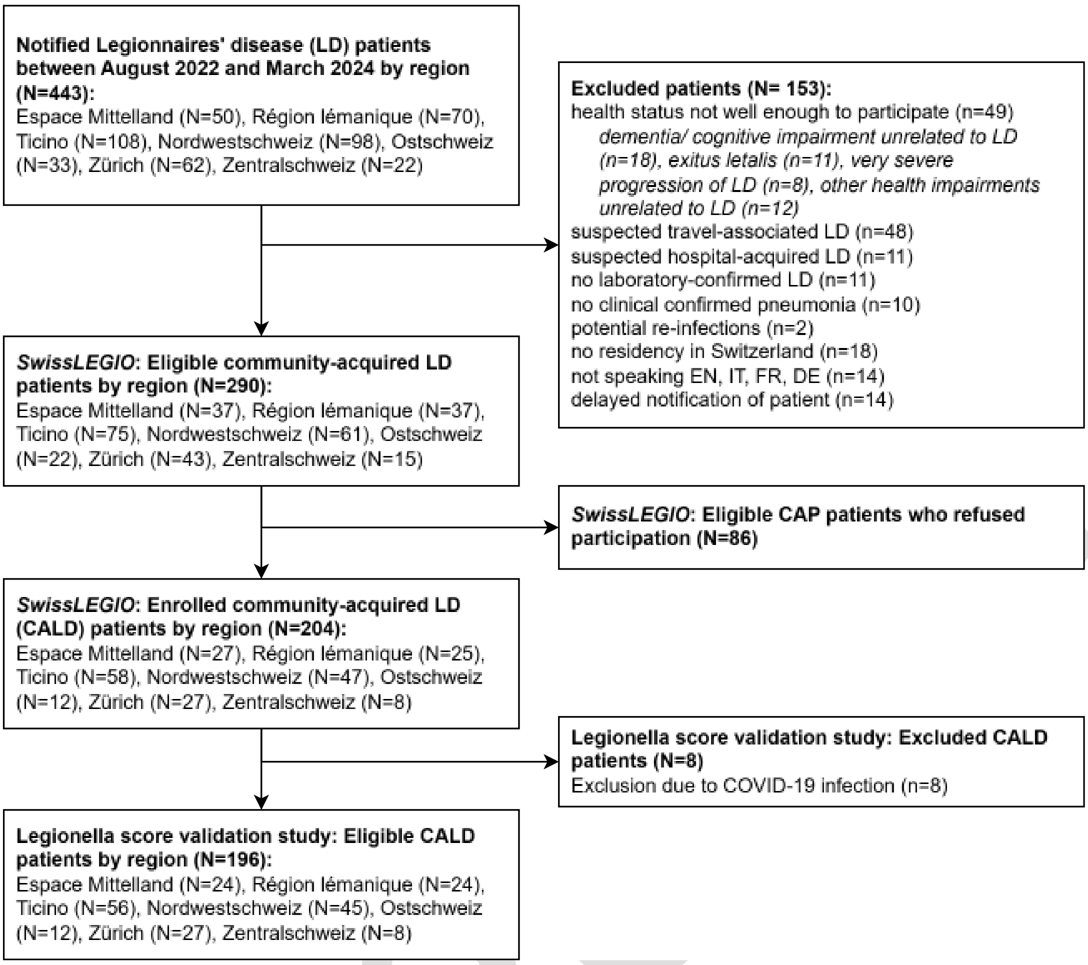

**SFigure 1** Flowchart showing the recruitment and inclusion of community-acquired LD (CALD) patients into the *SwissLEGIO* parent study and the score validation study.

**STable 1** Aetiology of pneumonia in Legionella test-negative CAP patients.

| | **Legionella test-negative CAP (n=196)** |
|---|---|
| Pathogen not known(%) | 124 (63.27%) |
| Pathogen known* | *n= 72* |
| *Streptococcus pneumoniae* | 29 (40.3) |
| *Mycoplasma pneumoniae* | 4 ( 5.6) |
| *Haemophilus influenzae* | 9 (12.5) |
| *Staphylococcus aureus* | 4 ( 5.6) |
| *Klebsiella pneumoniae* | 2 ( 2.8) |
| *Pseudomonas aeruginosa* | 7 ( 9.7) |
| Influenza-A | 9 (12.5) |
| Influenza-B | 1 ( 1.4) |
| Rhinovirus | 4 ( 5.6) |
| Respiratory-Syncytial-Virus (RSV) | 1 ( 1.4) |
| Parainfluenza | 3 ( 4.2) |
| Pneumocystis jirovecii | 3 ( 4.2) |
| Other | 13 (18.1) |

* Coinfections were reported for 13 (18.1%) patients:
S. *pneumoniae* and *H. influenzae;* S. *pneumoniae, H. influenzae* and *S.aureus;* S. *pneumoniae* and *S.aureus;* S. *pneumoniae, S.aureus* and *Influenza A;* S. *pneumoniae* and *RSV;* S. *pneumoniae* and other (2x)*; H. influenzae* and *S.aureus, ; H. influenzae, S.aureus* and *P. aeruginosa, ; H. influenzae* and *P. aeruginosa,; P. aeruginosa* and other (2x*); Influenza-B* and other

**STable 2** Comparison of patient characteristics of our non-LD CAP group with patient characteristics of other CAP studies from the US (Bellew *et al.*), Switzerland (Corridori *et al*.), and a multicentre study with centres in Europe, South Africa and the US (Haubitz *et al.)*

| | **non-LD CAP (n=196)** | **Non-LD CAP Bellew *et al.* (n=1909)** | **CAP* Luthi-Corridori *et al.* (n=254)** | **Non-LD CAP Haubitz *et al.* (n=1902)** |
|---|---|---|---|---|
| **Patient characteristics** | | | | |
| Male | 126 (64.3) | 932 (48.8) | 131 (51.6) | 1127 (59.3) |
| Age (years) (median [IQR]) | 72 [56-81] | 57 [46-71] | 78 [66–85] | 73 [58-82] |
| Current smoker | 50 (32.5) | 494 (25.9) | 33 (25.0) | - |
| **Comorbidities** | | | | |
| Heart failure | 38 (19.4) | 357 (18.7) | - | 340 (17.9) |
| COPD | 44 (22.4) | 449 (23.5) | 56 (22.0) | 473 (24.9) |
| Cerebrovascular diseases | 35 (17.9) | - | - | 295 (15.5) |
| Chronic kidney disease | 35 (17.9) | 263 (13.8) | 34 (13.4) | 215 (11.3) |
| Malignancies | 65 (33.2) | 458 (24.0) | 21 (8.3) | 245 (12.9) |
| Immunosuppression | 46 (23.5) | 319 (16.7) | NA | - |
| **Clinical manifestation at admission** | | | | |
| Fever > 38°C | 69 (35.6) | 486 (25.5) | - | |
| Body temperature (median [IQR]) | 37.2 [36.8-38.5] | - | 37.70 (±0.98) | 37.7 (36.7-38.3) |
| Productive cough | 93 (53.4) | - | - | 671 (35.3%) |
| Headache | 41 (20.9) | 875 (45.8) | - | - |
| Confusion | 17 (9.7) | 384 (20.1) | - | - |
| Diarrhoea | 29 (14.8) | 386 (20.2) | - | - |
| Nausea or Emesis | 30 (15.3) | 665 (34.8) | - | - |
| **Laboratory values at admission** | | | | |
| CRP, mg/L (median [IQR]) | 120.0 [50.3-211.3] | - | 129 (54.5-222.0) | 122 (45-235) |
| Procalcitonin, µg/L (median [IQR]) | 0.26 [0.11-1.63] | - | 0.29 (0.10, 0.91) | 0.44 (0.15-2.75) |
| Leukocyte count, x$10^9$/L (median [IQR]) | 12.10 [8.48-16.40] | - | 12.0 (8.9, 15.5) | - |
| Platelet count, x$10^9$/L (median [IQR]) | 231 [174-308] | - | - | 205 (150-235) |
| Lactate dehydrogenase, U/L (median [IQR]) | 229 [198-287] | - | - | 360 (274-475) |
| Sodium, mmol/L (median [IQR]) | 136 [133-139] | - | - | 137 (134-140) |
| Oxygen saturation <90% | 71 (36.8) | - | 38 (17.2) | - |
| **Disease progression and outcomes** | | | | |
| Length of stay (days, median [IQR]) | 7 [5- 11] | - | 6.5 (5–9) | 9 (6-11) |
| ICU admission | 40 (20.4) | 415 (21.7) | 24 (9.4) | 253 (13.3) |

* including 7 CALD cases, the study excluded immunocompromised patients (35/404) screened patients were immunocompromised)

#### Additional study results: Score validation

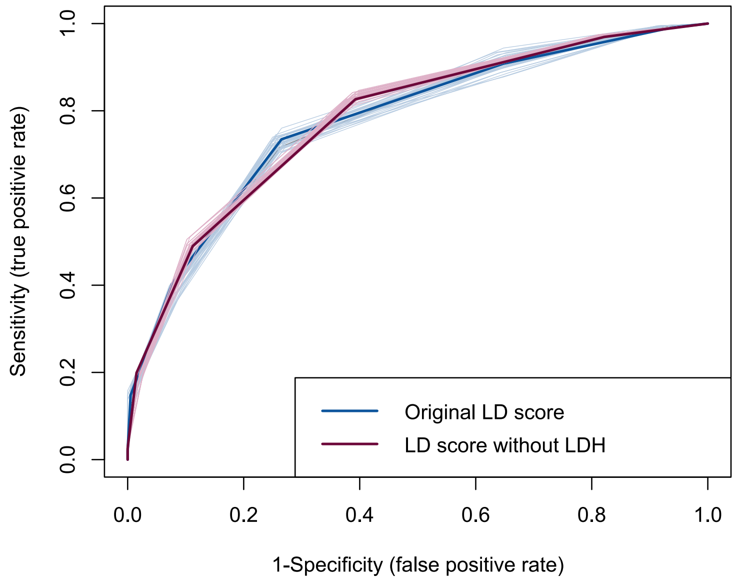

**SFigure 2** Receiver operating characteristic (ROC) curves for the original score by Fiumefreddo *et al.* and for the score when LDH was omitted. The two curves are very similar, indicating comparable predictive performance. Results from all 40 imputed datasets are shown, with one representative curve highlighted for improved readability.

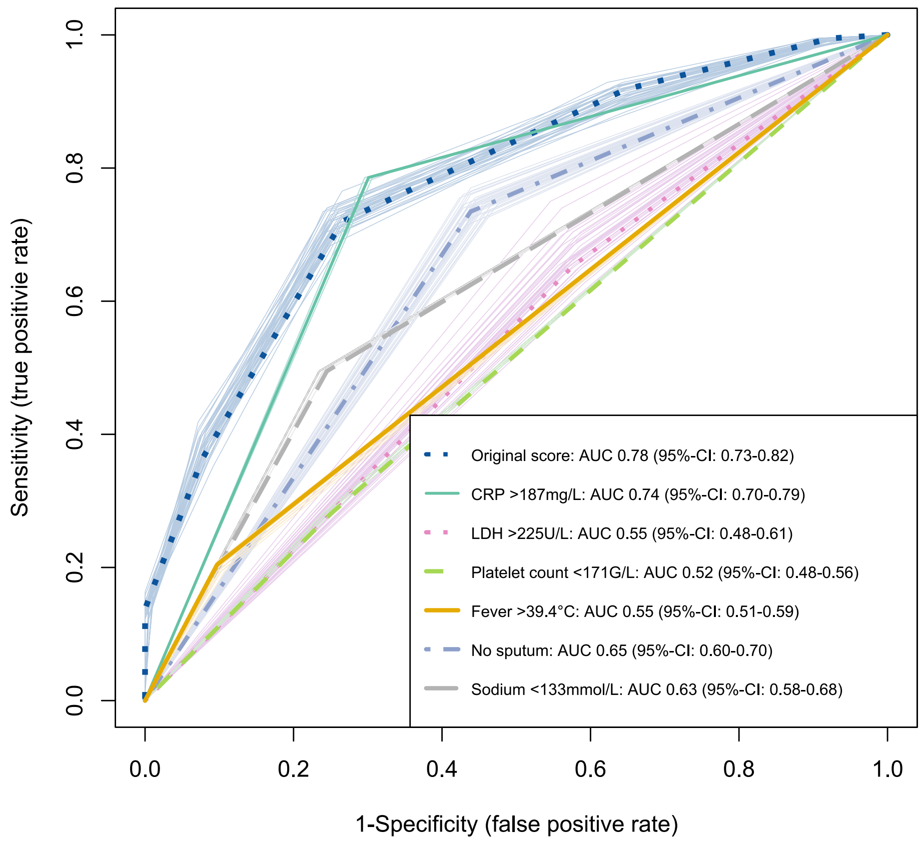

**SFigure 3** Receiver operating characteristic (ROC) curves for the individual (dichotomised) predictors and the original score by Fiumefreddo *et al.*. Results from all 40 imputed datasets are shown, with one representative curve highlighted for improved readability.

**STable 3** Estimated optimal cut-offs for the continuous predictors of the Legionella score proposed by Fiumefreddo *et al.*, calculated using Youden's index.

| Score predictors | Original cut-offs | Optimal cut-offs (95% CI) |
|---|---|---|
| No sputum (no cough/ dry cough) | - | - |
| **Fever (°C)** | **>39.4** | **37.4 (36.9 - 37.8)** |
| Sodium (Hyponatremia, mmol/L) | <133 | 134 (133-136) |
| Lactate dehydrogenase (LDH, U/l) | >225 | 264 (185-342) |
| C-reactive protein (CRP, mg/L) | >187 | 157 (95- 220) |
| Platelet count ($x10^9$/L) | <171 | 217 (152-281) |

Preprint

#### Additional study results: Score update

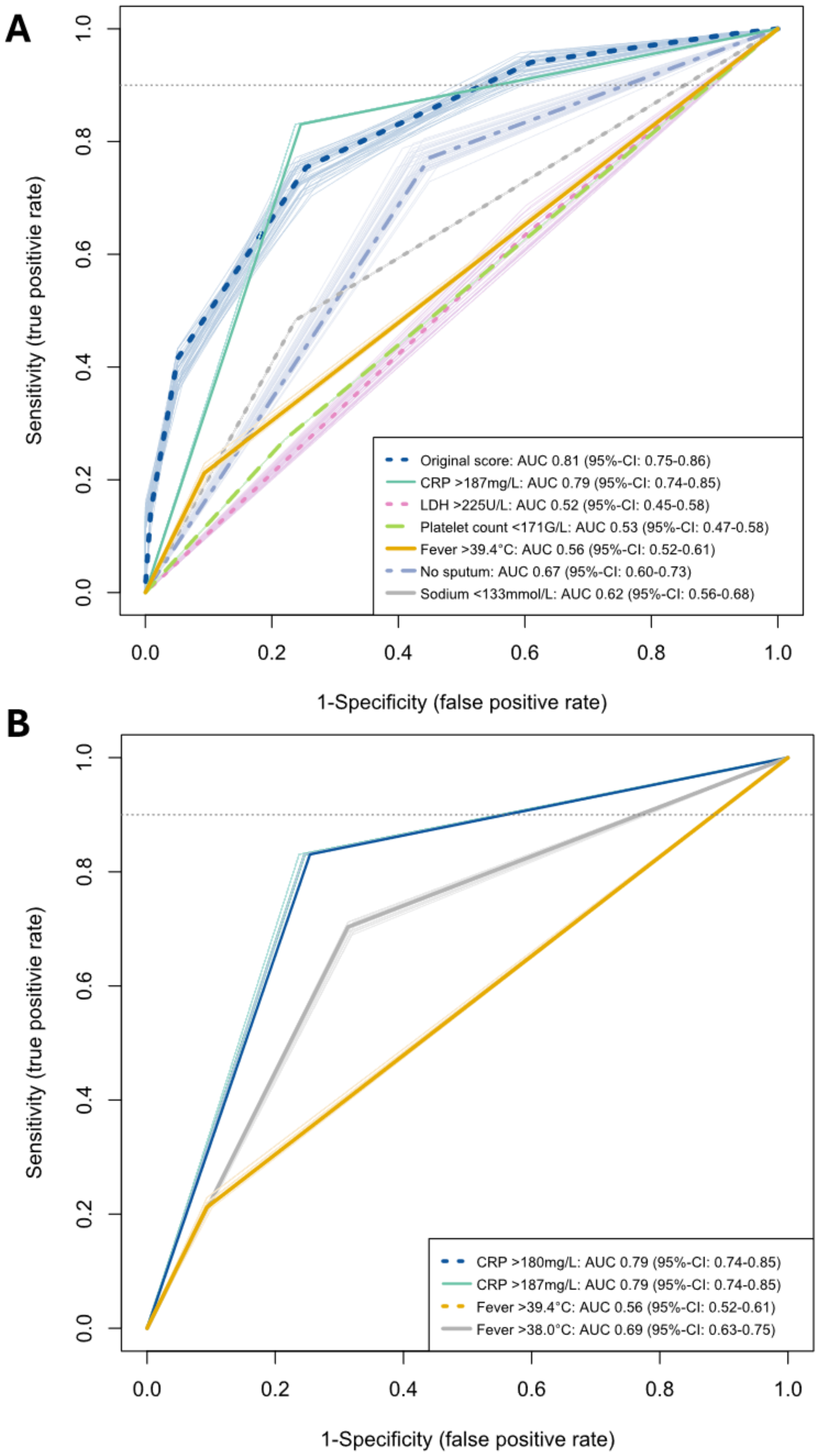

**SFigure 4** Receiver operating characteristic (ROC) curves for individual predictors (development data set, n=236). Results from all 40 imputed datasets are shown, with one representative curve highlighted for improved readability. A: individual predictors plus original score by Fiumefreddo *et al.*. B: Comparison old vs new cut-off for fever and CRP.

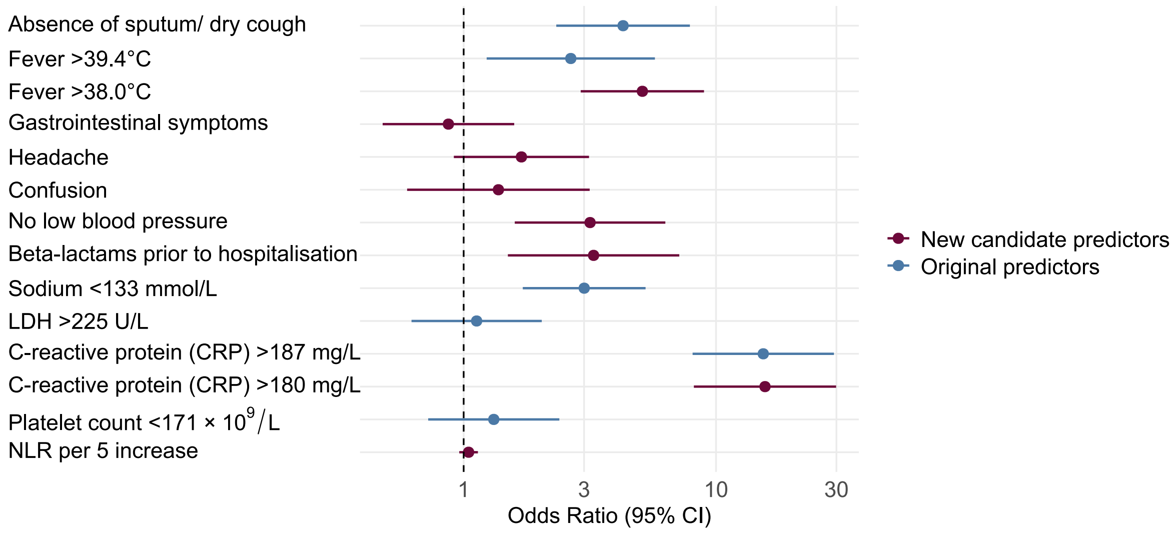

**SFigure 5** Results of the univariable regression analysis (development data set, n=236). No low blood pressure: >90/≥60 mmHg.

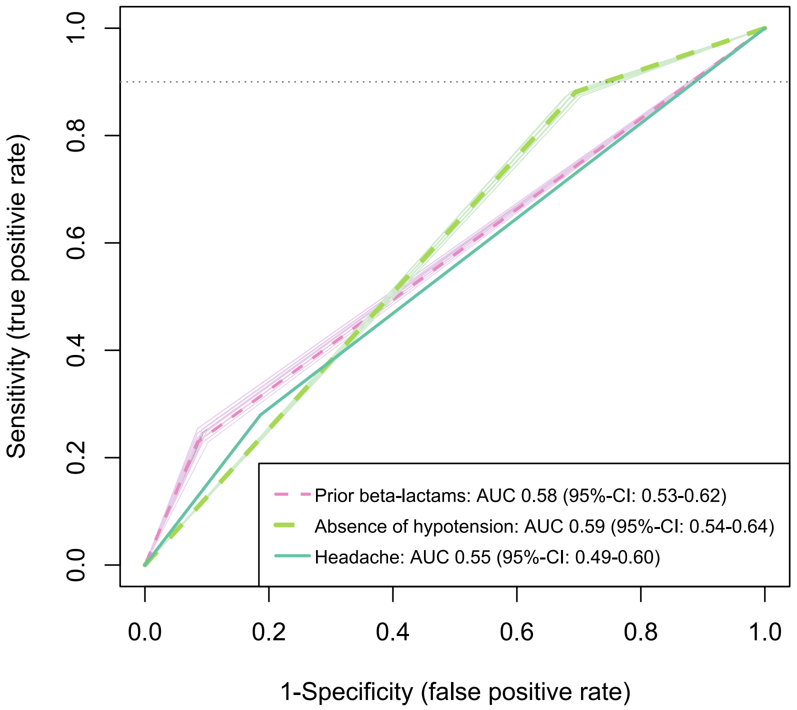

**SFigure 6** Receiver operating characteristic (ROC) curves for individual new candidate predictors (development data set, n=236). Results from all 40 imputed datasets are shown, with one representative curve highlighted for improved readability.

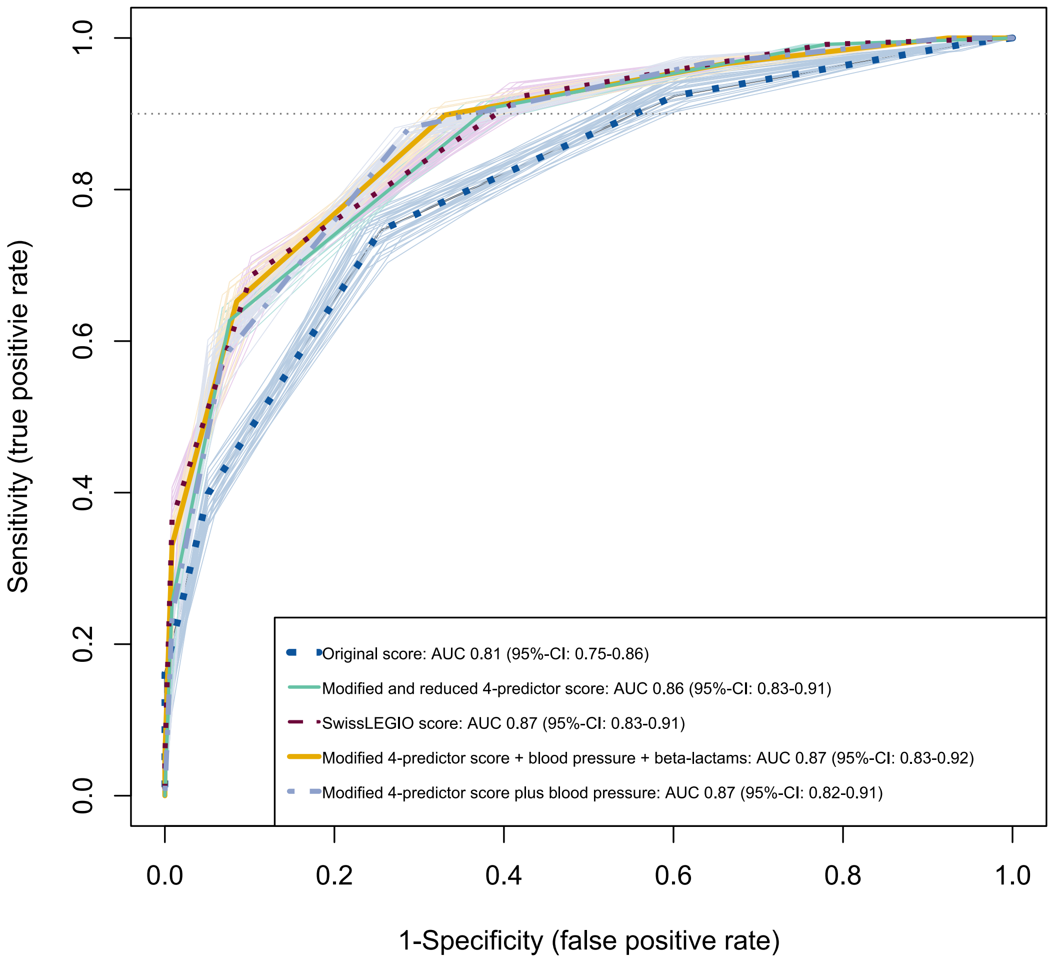

**SFigure 7** Receiver operating characteristic (ROC) curves for selected score specifications (development data set, n=236). Results from all 40 imputed datasets are shown, with one representative curve highlighted for improved readability. Original score as defined by Fiumefreddo *et al.*; modified and reduced 4-predictor score (CRP >180 mg/L, sodium <133 mmol/L, no/dry cough, and fever >38.0°C), *SwissLEGIO* score (CRP >180 mg/L, sodium <133 mmol/L, no/dry cough, fever >38.0°C, and prior β-lactam therapy).

**STable 4** Sensitivity and specificity for all score cut-offs for the original Fiumefreddo score, the modified and reduced 4-predictor score (CRP > 180 mg/L, sodium, no/dry cough, fever >38 °C), and the *SwissLEGIO* score (CRP > 180 mg/, sodium, no/dry cough, fever >38.0°C, prior β-lactam therapy) based on the development data set (n=236).

| Score cut-off | Original score Fiumefreddo *et al.* | | Modified, reduced 4-predictor score | | SwissLEGIO score | |
|---|---|---|---|---|---|---|
| | Sensitivity % (95%-CI) | Specificity % (95%-CI) | Sensitivity % (95%-CI) | Specificity % (95%-CI) | Sensitivity % (95%-CI) | Specificity % (95%-CI) |
| ≥0 | 100.0 (100.0–100.0) | 0.0 (0.0–0.0) | 100.0 (100.0–100.0) | 0.0 (0.0–0.0) | 100.0 (100.0–100.0) | 0.0 (0.0–0.0) |
| ≥1 | 99.1 (97.3–100.0) | 6.7 (1.8–11.7) | 99.2 (97.5–100.0) | 22.9 (15.1–30.7) | 99.2 (97.5–100.0) | 21.0 (13.5–28.5) |
| **≥2** | **93.3 (87.7–98.8)** | **40.0 (30.7–49.3)** | **90.6 (85.1–96.1)** | **61.8 (52.7–70.9)** | **92.3 (87.2–97.4)** | **58.1 (49.0–67.2)** |
| ≥3 | 74.0 (65.2–82.8) | 75.5 (67.3–83.7) | 63.0 (53.9–72.2) | 92.7 (87.9–97.5) | 68.7 (60.1–77.4) | 89.5 (83.7–95.2) |
| ≥4 | 39.2 (29.5–48.9) | 94.8 (90.8–98.9) | 26.1 (17.9–34.4) | 98.9 (97.0–100.0) | 38.3 (29.3–47.3) | 98.9 (97.0–100.0) |
| ≥5 | 13.5 (6.6–20.3) | 99.8 (98.7–100.0) | *NA* | *NA* | 4.7 (0.8–8.6) | 100.0 (100.0–100.0) |
| ≥6 | 1.0 (0.0–3.1) | 100.0 (100.0–100.0) | *NA* | *NA* | *NA* | *NA* |

**STable 5** Sensitivity and specificity for all score cut-offs for the original Fiumefreddo score, the modified and reduced 4-predictor score (CRP > 180 mg/L, sodium, no/dry cough, fever >38 °C), and the *SwissLEGIO* score (CRP > 180 mg/, sodium, no/dry cough, fever >38.0°C, prior β-lactam therapy) based on the validation data set (n=156).

| Score cut-off | Original score Fiumefreddo *et al.* | | Modified, reduced 4-predictor score | | SwissLEGIO score | |
|---|---|---|---|---|---|---|
| | Sensitivity % (95%-CI) | Specificity % (95%-CI) | Sensitivity % (95%-CI) | Specificity % (95%-CI) | Sensitivity % (95%-CI) | Specificity % (95%-CI) |
| ≥0 | 100.0 (100.0–100.0) | 0.0 (0.0–0.0) | 100.0 (100.0–100.0) | 0.0 (0.0–0.0) | 100.0 (100.0–100.0) | 0.0 (0.0–0.0) |
| ≥1 | 98.2 (94.2–100.0) | 7.9 (1.4–14.5) | 97.3 (93.6–100.0) | 14.2 (6.1–22.3) | 96.6 (92.1–100.0) | 8.8 (2.1–15.4) |
| **≥2** | **86.0 (76.6–95.5)** | **28.8 (17.9–39.8)** | **85.0 (76.9–93.1)** | **49.6 (37.8–61.3)** | **87.9 (79.9–95.9)** | **46.0 (34.0–57.9)** |
| ≥3 | 62.9 (49.9–75.9) | 73.5 (63.0–84.1) | 66.7 (55.9–77.5) | 80.5 (71.0–90.1) | 68.2 (56.9–79.5) | 74.3 (63.9–84.7) |
| ≥4 | 30.2 (18.2–42.3) | 88.4 (80.8–96.0) | 24.3 (14.5–34.2) | 97.5 (93.1–100.0) | 32.5 (21.2–43.7) | 95.2 (89.9–100.0) |
| ≥5 | 11.5 (3.4–19.7) | 99.8 (98.6–100.0) | *NA* | *NA* | 4.6 (0.0–9.7) | 100.0 (100.0–100.0) |
| ≥6 | 2.8 (0.0–6.9) | 100.0 (100.0–100.0) | *NA* | *NA* | *NA* | *NA* |

**STable 6** Positive predictive value (PPV) and negative predictive value (NPV) for selected CALD prevalences for the original Fiumefreddo score, cut-off ≥2

| Prevalence (%) | PPV % (95%-CI) | | NPV % (95%-CI) | |
|---|---|---|---|---|
| | Development data set (n=236) | Validation data set (n=156) | Development data set (n=236) | Validation data set (n=156) |
| 2 | 3.1 (2.5–3.8) | 2.4 (1.9–3.1) | 99.7 (99.2–100.0) | 99.0 (97.4–99.8) |
| 4 | 6.1 (5.0–7.5) | 4.8 (3.7–6.2) | 99.3 (98.4–99.9) | 98.0 (94.8–99.5) |
| 7 | 10.5 (8.7–12.8) | 8.3 (6.6–10.7) | 98.8 (97.1–99.8) | 96.5 (91.0–99.2) |
| 10 | 14.7 (12.3–17.8) | 11.8 (9.4–15.0) | 98.2 (95.8–99.7) | 94.9 (87.3–98.8) |

**STable 7** Positive predictive value (PPV) and negative predictive value (NPV) for selected CALD prevalences for the modified and reduced 4-predictor score (CRP > 180 mg/L, sodium <133 mmol/L, no/dry cough, fever >38 °C), cut-off ≥2.

| Prevalence (%) | PPV % (95%-CI) | | NPV % (95%-CI) | |
|---|---|---|---|---|
| | Development data set (n=236) | Validation data set (n=) | Development data set (n=236) | Validation data set (n=) |
| 2 | 4.6 (3.5–6.3) | 3.3 (2.5–4.7) | 99.7 (99.4–99.9) | 99.4 (98.8–99.8) |
| 4 | 9.0 (7.0–12.1) | 6.6 (4.9–9.1) | 99.4 (98.8–99.8) | 98.8 (97.5–99.5) |
| 7 | 15.2 (11.9–19.9) | 11.2 (8.5–15.3) | 98.9 (97.9–99.6) | 97.8 (95.6–99.2) |
| 10 | 20.9 (16.7–26.9) | 15.8 (12.1–21.1) | 98.3 (97.0–99.4) | 96.8 (93.6–98.8) |

**STable 8** Positive predictive value (PPV) and negative predictive value (NPV) for selected CALD prevalences for the *SwissLEGIO* score (CRP > 180 mg/, sodium, no/dry cough, fever >38.0°C, prior β-lactam therapy), cut-off ≥2

| Prevalence (%) | PPV % (95%-CI) | | NPV % (95%-CI) | |
|---|---|---|---|---|
| | Development data set (n=236) | Validation data set (n=) | Development data set (n=236) | Validation data set (n=) |
| 2 | 4.3 (3.4–5.7) | 3.2 (2.4–4.4) | 99.7 (99.5–99.9) | 99.5 (98.8–99.9) |
| 4 | 8.4 (6.6–11.0) | 6.3 (4.8–8.7) | 99.5 (98.9–99.8) | 98.9 (97.6–99.7) |
| 7 | 14.2 (11.4–18.3) | 10.9 (8.4–14.7) | 99.0 (98.1–99.7) | 98.1 (95.8–99.5) |
| 10 | 19.7 (15.9–24.8) | 15.3 (11.9–20.2) | 98.5 (97.2–99.6) | 97.2 (93.9–99.2) |

### Additional information: Study methods

#### *Formula positive and negative predictive values*

$$PPV = \frac{sensitivity * prevalence}{(sensitivity * prevalence) + ((1 - specificity) * (1 - prevalence))}$$

$$NPV = \frac{specificity * (1 - \ prevalence)}{(specificity * (1 - prevalence)) + ((1 - sensitivity) * prevalence)}$$

[*] *Prevalence: pre-test probability of the disease (CALD) in the population being tested (all CAP patients)*

*Numer-needed-to-test (NNT)= 1/PPV*

#### *Candidate predictors for score update*

**STable 9** Candidate predictors from the literature. In bold (blue) predictors that we considered for the update of the score in the univariable model.

| Variable | Example reference | Missing in our data set (%) |
|---|---|---|
| Absence of certain comorbidities: COPD, heart failure | (1-5) | 0.0 |
| Elevated Procalcitonin | (1, 2, 6) | 66.3 |
| Younger age | (1-4, 7) | 0 |
| **No hypotension** | (7, 8) | **2.6** |
| **GI symptoms** | (3, 5, 6, 8-10) | **0** |
| **Neutrophils/ WBC (NLR)** | (6) | **11.2** |
| Smoking | (3, 4, 7, 9, 11) | 11.5 |
| **Headache** | (3, 5, 7-10) | **0** |
| **Confusion** | (5, 8, 10, 11) | **7.4** |
| Male sex | (3, 4, 11) | 0 |
| **Previous beta-lactam therapy** | (1, 7-9) | **5.1** |

Preprint
